# Informed Consent in an International Trial of Stress Ulcer Prophylaxis - Patterns and Predictors: A Protocol and Statistical Analysis Plan

**DOI:** 10.1101/2025.07.17.25330833

**Authors:** Diane Heels-Ansdell, Sangeeta Mehta, Karen E. A. Burns, Nicole Zytaruk, France Clarke, Miranda Hardie, Simon Finfer, Deborah Cook

**Author notes:** **Correspondence** Dr. DJ Cook, Academic Critical Care Office D176, St. Joseph’s Healthcare Hamilton, 50 Charlton Avenue East, Hamilton, Ontario, Canada L9H 4A6. for the REVISE Research Coordinators, REVISE Investigators, Canadian Critical Care Trials Group and the Australian and New Zealand Intensive Care Society Clinical Trials Group.

## Abstract

**Background:** Informed consent rates are inconsistently incorporated in trial reports, and literature on consent patterns and predictions is sparse, particularly in the field of critical care.

**Objective:** The overall objective of this study is to describe the patterns and predictors of consent rates in REVISE. The specific aims are to analyze the consent models used, consent rates, participants in the consent encounter, reasons for declined consent, and factors associated with obtaining consent.

**Methods:** This is a pre-planned secondary study of the REVISE Trial (NCT03374800) which compared pantoprazole to placebo on the outcome of clinically important upper gastrointestinal bleeding among invasively ventilated patients in the intensive care unit (ICU). Research ethics committees approved the protocol in all jurisdictions. Research personnel prospectively collected standardized data for each consent encounter, including the consent model (*a priori*, deferred, or opt-out), the role of the individual who provided or declined consent (patient, SDM, other), and the reasons for declined consent, the role of the individual who requested consent (research coordinator, site investigator, ICU physician) and the consent encounter method (in person or via telephone). When consent was provided and then later revoked, who revoked consent (patient, SDM, other) and timing (in ICU, in hospital, post hospital), as well as details about permission for data retention were collected. In this study, consent rates will be calculated across REVISE sites, and in relation to the COVID-19 pandemic. We will also calculate the consent rates for *a priori* and deferred consent models, the timing from deferred recruitment to consent provided or declined, which personnel requested consent (research coordinator, site investigator, ICU physicians, other), and who engaged in the consent encounter for each consent model (patient, SDM, other). Multilevel logistic regression analysis will be conducted to evaluate variables independently associated with consent including additional site and staff variables.

**Results:** By analyzing the frequency and impact of consent models, and consent encounters with various stakeholders, results will highlight the acceptability of different approaches, and the impact of different approaches in this critical care trial.

**Conclusions:** This pre-planned retrospective sub-study using an international randomized controlled trial database will provide useful informed consent metrics and knowledge that is relevant to contemporary global trial conduct.

## INTRODUCTION

Consent to participate in research requires consent to be informed, voluntary, documented and ongoing (CIHR). Most critically ill patients are incapable of decision-making, such that substitute decision-makers (SDMs) are typically approached to consider research opportunities on their behalf (Burns). Through soliciting the consenting experiences of North American research coordinators who worked on an international thromboprophylaxis trial in the ICU setting (Cook), we previously developed guidance when approaching SDMs regarding the enrolment of critically ill patients into randomized trials in 3 phases: preparing for the consent encounter, during the consent encounter, and follow-up to the consent encounter. The 13 strategies we collated reinforce requirements for the informed consent outlined in existing legislation and highlight additional processes that may enhance the integrity of the consent process (Smith).

SDMs in the ICU may be particularly susceptible to therapeutic misconception, given the complexity of the clinical situation, along with high morbidity and mortality associated with critical illness. A Canadian survey of SDMs who agreed to have a relative participate in a critical care study reported being motivated by the potential benefit to the patient and altruism, whereas those who declined - while not generally opposed to research - were fearful of study-related harm or discomfort for the patient, and too anxious to consider research participation (Mehta). Informed consent rates are inconsistently incorporated in trial reports, and literature on consent patterns and predictions is sparse.

This is a protocol and statistical analysis plan for a retrospective analysis of the <u>R</u>e-<u>Ev</u>aluating the Inhibition of <u>S</u>tress <u>E</u>rosions (REVISE) Trial (NCT03374800). REVISE was an international stratified, concealed, blinded, parallel group randomized controlled trial evaluating the proton pump inhibitor pantoprazole compared to placebo for stress ulcer prophylaxis among 4821 critically ill patients undergoing mechanical ventilation (Cook). The primary efficacy outcome was clinically important upper gastrointestinal bleeding.

The overall objective of this study is to describe the patterns and predictors of consent rates in REVISE. The specific aims are to analyze the consent models used, consent rates, participants in the consent encounter, reasons for declined consent, and factors associated with obtaining consent.

## METHODS

### Design

This is a pre-planned secondary study of the REVISE Trial (NCT03374800). Adults ≥18 years old receiving invasive mechanical ventilation and expected to remain mechanically ventilated beyond the calendar day after randomization were eligible. The full trial protocol, statistical analysis plan and the results of the primary analysis have been published (Deane, Heels-Ansdell, Cook). Research ethics committees, regulators and/or local ICU leadership approved the launch of new participating sites during the COVID-19 pandemic based on capacity evaluation and public health decisions in each jurisdiction. Enrolment commenced on July 9, 2019 until October 30, 2023, spanning the COVID-19 pandemic, and was conducted in 8 countries.

Three approaches were used for trial participation in 68 centers: *a priori* consent, deferred consent, and an opt-out consent model. A priori consent involves obtaining agreement to participate in a consent encounter before enrolling a patient in a study. Deferred consent (also referred to as consent to continue) involves enrolling a patient in a study, then having a consent encounter with the patient or SDM to seek agreement. The opt-out approach involves enrolling an eligible patient, then informing the patient or SDM afterwards and allowing their subsequent withdrawal from continued participation if requested. The waived consent model involves enrolment but requires no explicit notification about study enrolment at any time.

### Data collection

In REVISE, research personnel prospectively collected standardized data at the time of each consent encounter. These data included the consent model (*a priori*, deferred, or opt-out), the role of the individual who provided or declined consent (patient, SDM, other), and the reasons for declined consent, as applicable. Research personnel also collected the role of the individual who requested consent (research coordinator, site investigator, ICU physician) and the consent encounter method (in person or via telephone), except for a priori declines. When consent was provided and then revoked at a later time, data on who revoked consent (patient, SDM, other) and timing (in ICU, in hospital, post hospital), as well as details about permission for data retention were collected.

After trial completion, in November-December 2024, participating sites completed a 1-page site questionnaire (Appendix 1). We requested information regarding site characteristics, including annual ICU admissions in 2023, number of ICU beds screened for REVISE, types of patients routinely admitted to their ICU, and teaching hospital status (not university-affiliated, limited teaching, or full teaching institutions). Research teams also self-reported the number of pages in their center’s REVISE consent form, research staff capacity, research staff trial-related experience, their center’s participation in randomized trials over the previous 5 years, whether their center is affiliated with a research institute, whether the center permitted co-enrolment of individual patients into more than one study, and any co-enrolment guidance followed (local policy, national guidelines, other).

### Statistical analysis

The protocol and statistical analysis plan for this secondary retrospective analysis were reviewed and approved by the Canadian Critical Care Trials Group. Characteristics of sites and consent encounters will be summarized using proportions or categorical data and mean (SD) or median (interquartile range), as appropriate. Data from the single site which used the opt-out model will be excluded from some of these analyses, as there was not a conventional consent encounter.

Consent rates will be calculated across REVISE sites, and in relation to the COVID-19 pandemic. We will define temporal categories as: pre pandemic = July 2019 to March 10, 2020; COVID-19 pandemic = March 11, 2020, to May 31, 2023, as per World Health Organization declared dates; and post pandemic = June 1, 2023 - Oct 30, 2023, when the last patient was randomized.

We will calculate the consent rates for *a priori* and deferred consent models, the timing from deferred recruitment to consent provided or declined, which personnel requested consent (research coordinator, site investigator, ICU physicians, other), and who engaged in the consent encounter for each consent model (patient, SDM, other).

Multilevel logistic regression analysis will be conducted to evaluate variables independently associated with consent. The final variables will be in the domains of center characteristics and consent encounter characteristics, and selected based on previously published studies, and through discussion among co-authors. We hypothesize that higher consent rates would be associated with deferred consent (vs. *a priori*), consent encounter with SDM (vs. patient), the COVID-19 pandemic period, larger ICUs, ICUs permitting co-enrolment, ICUs with more trial related experience, and research staff with greater experience. Analyses will be performed using SAS software, version 9.4.

## RESULTS

This REVISE consent processes and predictors study will analyze the frequency and impact of consent models, and consent encounters with various stakeholders. Findings will highlight the acceptability of different approaches, and the impact of allowing deferred consent in a trial that was largely implemented during the COVID-19 pandemic.

## DISCUSSION

In summary, this pre-planned retrospective sub-study using a contemporary international randomized controlled trial database will provide useful informed consent metrics and knowledge that is relevant to global trial conduct.

Strengths of this study will include the large number of participants enrolled in REVISE internationally, and prospective collection of detailed data pertaining to consent encounters and participants. However, patterns and predictors of consent rates may differ in other trials testing difference interventions, and involving greater risk, in different jurisdictions.

## Data Availability

Not applicable - Protocol Manuscript

